# Unsupervised Phenotypic Stratification of 22q11.2 Deletion Syndrome for Improved Clinical Screening

**DOI:** 10.64898/2026.10.30.26366765

**Authors:** Laís Azevedo Soares, Deborah Domeneghetti de Francisco, Vera Lúcia Gil-da-Silva-Lopes, Guilherme Palermo Coelho, Ana Estela Antunes da Silva

## Abstract

**Objective:** 22q11.2 deletion syndrome (22q11DS) is the leading genetic cause of palatal anomalies, presenting a heterogeneous spectrum of clinical manifestations that complicates early diagnosis. No study has systematically investigated the co-occurrence of symptoms spanning cardiovascular, neuropsychiatric, craniofacial, immune, endocrine, renal, and familial domains within a single cohort using unsupervised learning methods for 22q11DS stratification. This study proposes a pipeline to identify subgroups and associated symptom patterns from binary clinical data, aiming to improve screening and referral for genetic testing.

**Design:** Cross-sectional, retrospective study comparing three unsupervised clustering frameworks— K-medoids, DBSCAN, and Latent Class Analysis (LCA)—followed by association rule mining applied to individuals with confirmed deletion.

**Setting:** Brazilian Database of Craniofacial Anomalies (BBAC), a national clinical registry.

**Participants:** A cohort of 297 individuals with suspected 22q11DS.

**Main Outcome Measures:** Cluster separability (silhouette coefficient), characterization of phenotypic subgroups, and sensitivity/specificity of the current screening protocol stratified by subgroup.

**Results:** LCA achieved the highest cluster separability (silhouette = 0.165), yielding four distinct subgroups: Cardiac-predominant (C0), Craniofacial-palatal (C1), Neurodevelopmental-psychiatric (C2), and Mild-craniofacial (C3). Cluster coherence was confirmed by medical specialists. Quantitative protocol assessment revealed adequate overall sensitivity, with markedly lower performance for C2 and C3, identifying profiles at higher risk of being missed by current screening criteria.

**Conclusions:** These findings provide a data-driven basis for recalibrating symptom weights and referral thresholds, improving sensitivity for underperforming subgroups while reducing unnecessary referrals, optimizing the balance between diagnostic coverage and the financial burden of genetic testing.

## 1 Introduction

Anomalies are present in approximately 3% to 5% of all live births and include craniofacial mal-formations such as orofacial clefts, craniosynostosis, mandibulofacial dysostosis, and macrosomia among the most common alterations.^1, 2^ The 22q11.2 deletion is the leading genetic cause of palatal anomalies and the second most frequent cause of developmental delay associated with congenital heart defects, in addition to being the most common microdeletion syndrome.^3–5^ It is a rare disease, with an estimated prevalence ranging from 1:3,000 to 1:6,000 live births, whose diagnosis often depends on costly genetic testing.^6, 7^ Therefore, assembling a large cohort of cases for clinical data analysis becomes challenging.

The 22q11.2 deletion syndrome is highly heterogeneous, with manifestations ranging from subtle to complex, even within members of the same family. This clinical variability complicates diagnosis, particularly when signs are not evaluated collectively.^6, 8^ The challenge is further amplified by phenotypic overlap with other conditions such as CHARGE, Noonan and Wiedemann-Steiner syndromes,^9–11^ and by the recognition that not all observed manifestations are necessarily attributable to the deletion itself — additional genetic or environmental influences, including epigenetic modifications and mosaicism,^8, 10^ may independently affect gene expression and contribute to the observed phenotypic variability. Given this complexity, final diagnosis relies on genetic testing, including FISH, MLPA, CMA, and Whole Exome Sequencing, each offering different levels of genomic resolution. Previous research indicates that, although CMA is roughly ten times more expensive than other approaches in the Brazilian public healthcare system, it still offers the most extensive genome-wide assessment.^5, 12^

In this context, Brazil’s Craniofacial Project (BCFP) was established in 2003 as a voluntary multicenter network aimed at generating scientific evidence on craniofacial anomalies,^13^ including the creation of the Brazilian Database on Craniofacial Anomalies (BBAC) for standardised population-based clinical data collection.^14^ Based on longitudinally collected data through the BBAC, a clinical guideline was developed to optimise decision-making in suspected 22q11.2 deletion cases, incorporating a hierarchical diagnostic approach in which individuals are first filtered based on clinical criteria, followed by FISH and MLPA as primary genetic screening tests, with CMA reserved for cases that remain under clinical suspicion following negative initial results.^12^ Individuals fulfilling the proposed screening protocol criteria were 3.82 times^12^ more likely to receive a confirmed 22q11.2 deletion diagnosis, with the complete hierarchical strategy estimated to generate savings of $175,135.13 in the Brazilian public health system (SUS) for each group of one thousand individuals.^12^ Nevertheless, diagnostic accuracy reached up to 34.7% depending on the combination of manifestations analysed, indicating that a substantial proportion of cases remains undetected, suggesting that data-driven approaches may help to capture phenotypic patterns that the current guideline has yet to formalise. From a machine learning standpoint, previous studies have applied unsupervised methods to characterise heterogeneity in 22q11.2 deletion syndrome. Using behavioural, morphological, and neuroimaging data from 50 paediatric individuals, k-means clustering identified distinct neurobehavioral subgroups.^15^ Hierarchical clustering applied to DNA methylation data revealed three epigenetically distinct subgroups within the deletion spectrum.^16^ Latent class analysis of cognitive profiles further demonstrated gradients of overall functioning rather than discrete phenotypic subtypes.^17^ Nevertheless, although these studies highlight the clinical heterogeneity of the syndrome and demonstrate the utility of clustering for subgroup identification, no previous study has conducted a comprehensive multivariate analysis combining cardiovascular, neuropsychiatric, craniofacial, immune, endocrine, renal, and familial features simultaneously. Furthermore, no systematic comparison of clustering frameworks has been conducted to identify the most suitable approach for binary clinical data in this population, nor have co-occurring symptom patterns been examined across these domains in conjunction with a quantitative assessment of the clinical screening protocol currently used.^18^

Hence, this study aims to investigate the clinical profiles of individuals using BBAC collected data^14^ in a cohort of 297 individuals evaluated for 22q11.2 deletion syndrome, including 94 confirmed positive cases. Clustering analysis was performed to identify subgroups, comparing three different unsupervised methods: K-Medoids (distance-based),^19^ DBSCAN (density-based),^20^ and Latent Class Analysis (probabilistic model-based).^21^ Subsequently, the best-performing technique was selected, and association rule mining was applied within each generated cluster to further characterise the clinical patterns of the individuals with confirmed deletion in each subgroup. To validate the clinical relevance of the resulting subgroups, the identified clusters and association rules were reviewed by medical specialists in clinical genetics and craniofacial anomalies, who assessed their coherence with established syndromic knowledge and current diagnostic practice.

The present study investigates whether clinically meaningful subgroups exist within the 22q11.2 deletion syndrome population based on co-occurring symptom patterns, and whether an unsupervised pipeline can elucidate these structures to support improved screening and genetic referral in a real clinical setting. To this end, this paper is organised as follows: Section 2 describes the dataset, preprocessing pipeline, clustering framework comparison, and association rule mining; Section 3 reports the identified latent classes, MCA visualisations, and their clinical feature profiles; Section 4 presents the medical specialist validation of the identified subgroups, their implications for the current craniofacial screening protocol, limitations, and future directions; Section 5 concludes the paper.

## 2 Methods and Procedures

The proposed method encompasses five main stages, as illustrated in Fig. 1. Unsupervised learning techniques are applied to uncover phenotypic patterns without relying on labelled outcomes, making them suitable for exploratory pattern discovery rather than classification. Clustering is used to partition individuals into groups based on inter-sample similarity, where the chosen distance metric determines which individuals converge into the same subgroup. Three clustering algorithms are systematically compared, as each computes similarity differently, potentially generating different groups.^22^ Since clustering does not incorporate clinical outcome information, association rule mining is applied as a complementary step. Association rules are mined within each cluster using all of its members, with confirmed deletion status fixed as the consequent, enabling the extraction of co-occurring symptom patterns associated with a positive diagnosis in each subgroup.

**Figure 1:**
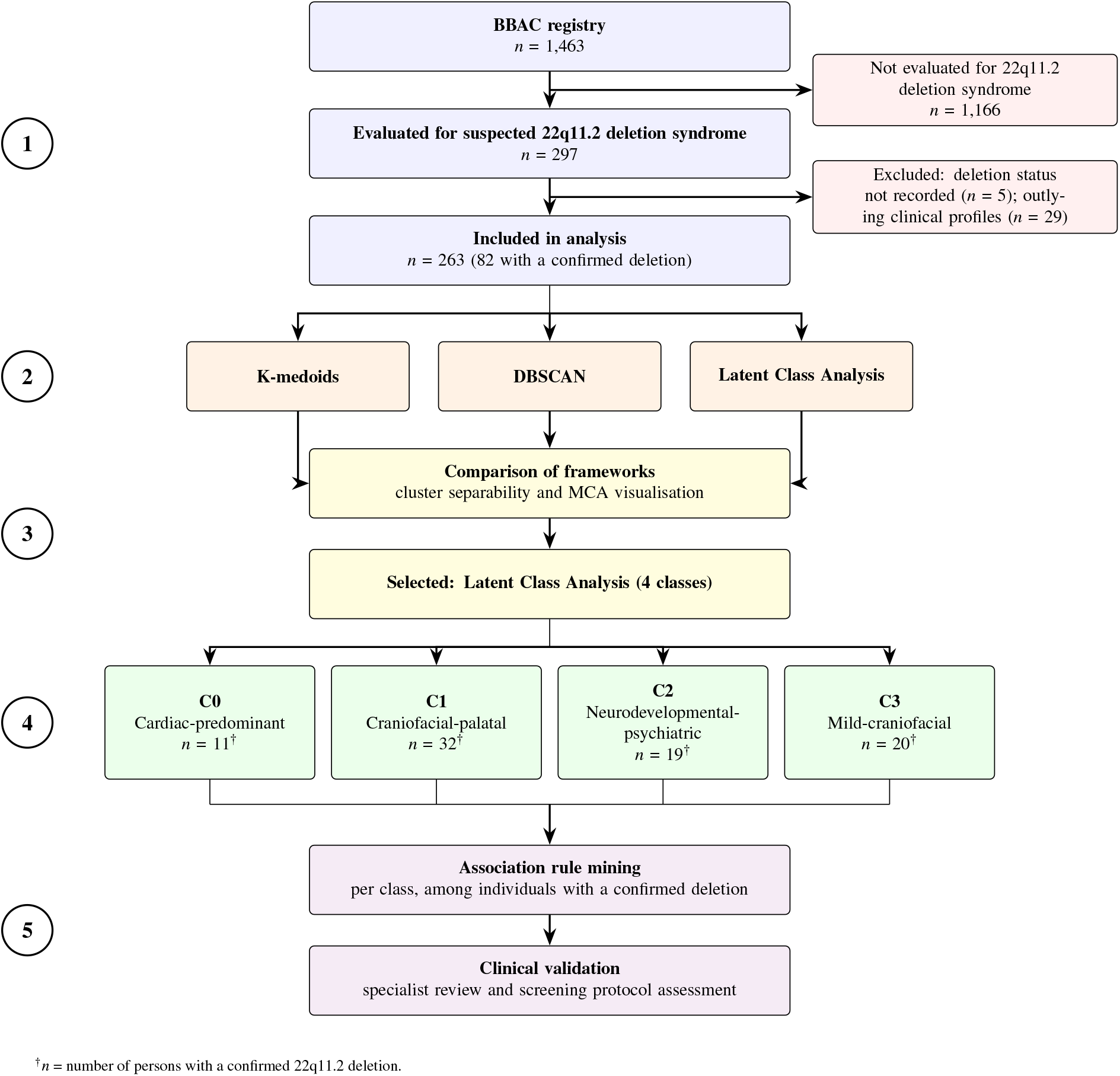
Analytical pipeline for phenotypic stratification of persons evaluated for 22q11.2 deletion syndrome (n = 1,463; BBAC registry). After exclusions, 263 participants (82 with a confirmed deletion) were analysed. Three unsupervised clustering frameworks (K-medoids, DBSCAN, Latent Class Analysis) were compared by cluster separability and MCA visualisation; Latent Class Analysis yielded four phenotypic classes (C0–C3). Association rule mining was applied per class, followed by clinical validation through specialist review.

### 2.1 Dataset Description

Data were collected through Brazil’s Craniofacial Project using the CranFlow software (INPI No. BR2015510005502), comprising records from genetic centres across Brazil under ethics approval (CAAE No. 85020018.8.0000.5404).^14^ It covers 1,463 individuals with different craniofacial anomalies, including 462 syndromic individuals and 408 attributes ranging from demographics and exam results to facial features, structural malformations, and clinical characteristics across a variety of anomalies. For this study, all individuals with suspicion of 22q11.2 deletion were selected (297 samples), covering 94 positive cases. Individuals in the database vary in age and sex, as shown in Fig.

2. The predominant age group was 5–9 years, with adults and infants represented at lower frequencies; 23 individuals (7%) had sex not recorded. This age distribution is significant to highlight, as younger individuals may not yet have manifested the full spectrum of syndromic features, potentially affecting the completeness of symptom profiles in this group.

**Figure 2:**
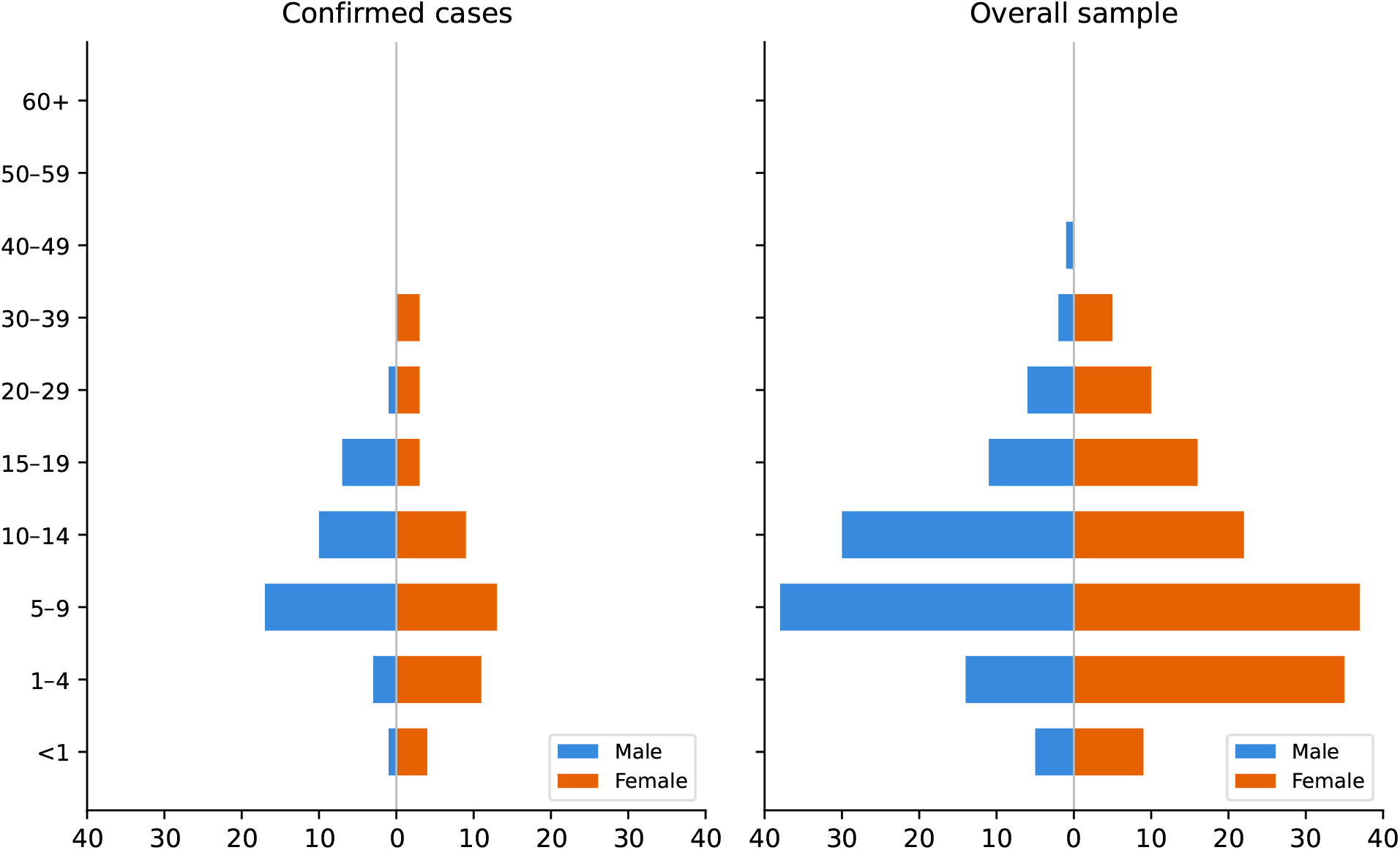
Age and sex distribution of the overall sample and confirmed cases.

### 2.2 Feature Selection and Filtering

Feature selection was performed by the responsible medical team based on the representative characteristics defined in the 22q11.2 screening protocol,^18^ resulting in 57 clinical features, which limited the inclusion of attributes associated with other craniofacial anomalies present in the dataset. Following feature engineering, a variance threshold of 0.01 was applied to remove near-zero variability features. The final dataset comprised 45 clinical features used as input for the clustering analysis, alongside two outcome variables retained separately for validation: *Deletion Positive* and *Meets Screening Criteria*, presented in Fig. 4.

**Figure 3:**
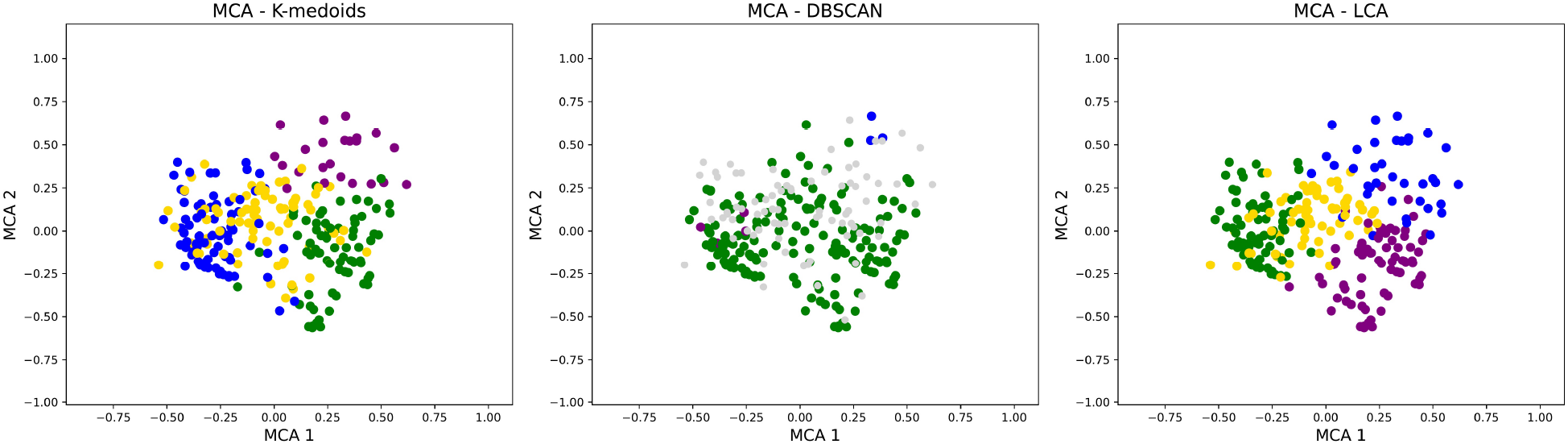
Multiple correspondence analysis (MCA) projections for the best parameter configuration of each clustering method.

**Figure 4:**
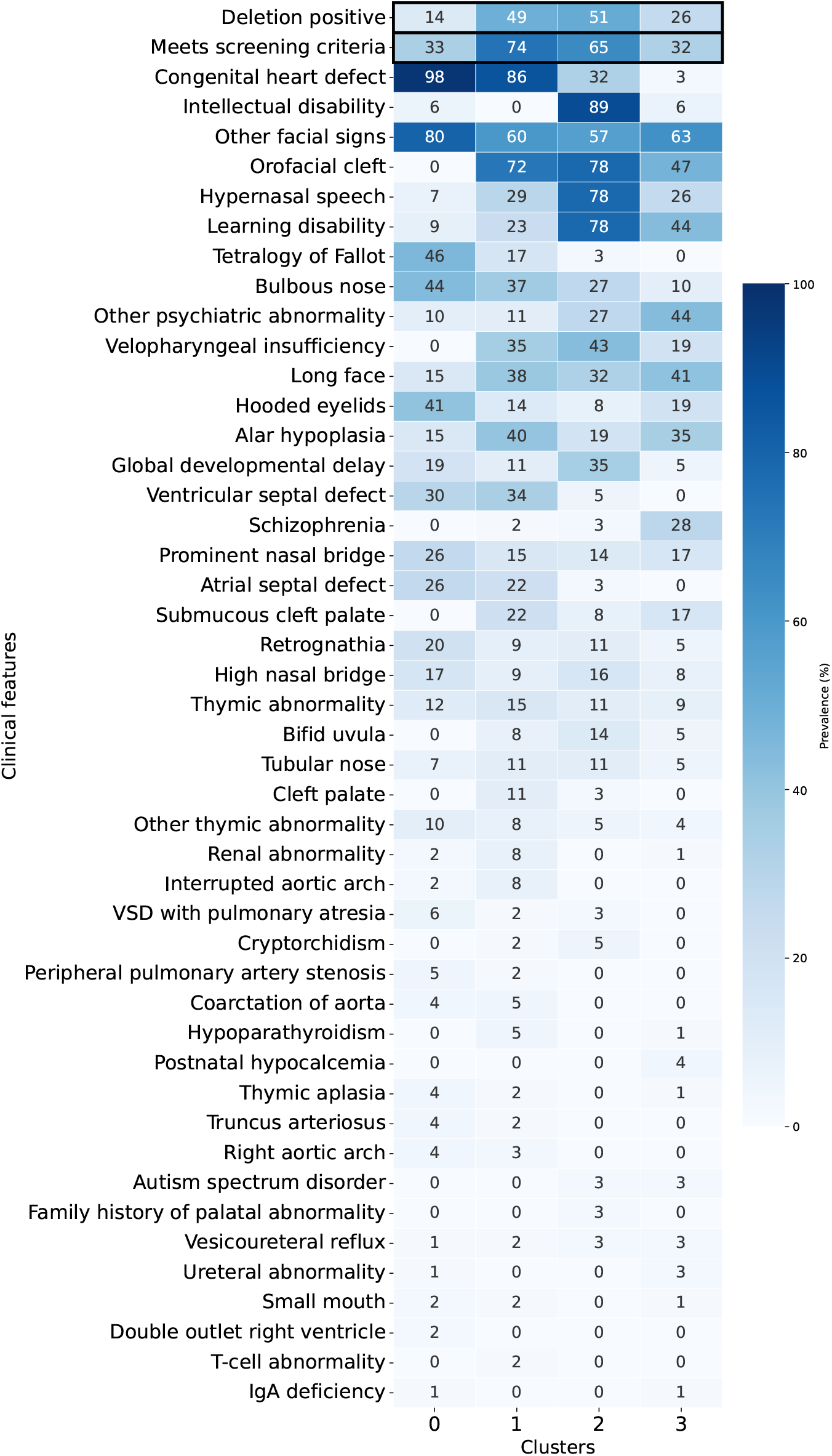
Heatmap of symptom prevalence by cluster. Each cell represents the percentage of individuals positive for a given clinical feature within each cluster, with darker shades of blue indicating higher prevalence. The first row displays the proportion of deletion-positive individuals, while the second row shows the proportion of individuals who met the protocol screening criteria.

### 2.3 Feature Engineering and Data Preprocessing

The distribution of each clinical attribute was analysed to assess missing data prevalence. Nine attributes exhibited a high proportion of missing values, however their exclusion was not feasible given their clinical relevance, and removing affected samples would have disproportionately reduced the number of individuals with a confirmed deletion. Attributes with more than two missing observations were therefore one-hot encoded, expanding each categorical variable into multiple binary support columns, whereby missing values are represented as a distinct binary indicator rather than treated as absence of the feature.^23^

Individuals with confirmed, typical, or atypical deletions were classified as positive, following the literature consensus.^24, 25^ Individuals with missing values for the primary outcome variable (*Deletion Positive*) were excluded from the analysis (*n*=5), as deletion status is required for cluster validation and screening protocol assessment, and its absence could introduce noise into the clustering procedure. To reduce redundancy and dimensionality, intellectual disability was defined as positive if any of the following sources reported a positive finding: formal neuropsychological evaluation, medical staff clinical assessment, or the general intellectual disability. Tetralogy of Fallot was binarized such that all positive presentations (classic, stenotic, or unspecified) were coded as positive, while absence of the condition was coded as negative.

The two validation attributes, *Deletion Positive* and *Meets Screening Criteria*, were excluded from the clustering procedure, resulting in a final feature space of 61 binary dimensions used for analysis. Given the binary nature of the data, conventional outlier detection methods are not directly applicable. Outlier removal, which consists of excluding observations that deviate substantially from the overall data distribution, was performed by computing the Euclidean distance of each observation from the global centroid, excluding observations beyond the 90th percentile. This threshold was chosen empirically by observing the effect of different thresholds on the Multiple Correspondence Analysis (MCA) projections (described in Section 2.5), selecting the value that removed only observations visibly distant from the main data distribution while minimising sample loss, given the inherent scarcity of clinical data in rare disease cohorts. Following removal, 29 samples were excluded — including 12 confirmed positive cases — reducing the positive subset from 94 to 82 cases.

### 2.4 Clustering paradigms

Clustering method selection was guided by the binary nature of the data and the features with potential outliers.^22^ Hence, three algorithmic frameworks were evaluated — distance-based, density-based, and probabilistic.

K-medoids is a distance-based clustering algorithm that partitions data into k clusters, using actual samples as cluster representatives (medoids) rather than computed centroids, an appropriate property for binary clinical data, where a mean point may not correspond to any real individual profile.^19^ The alternate optimisation method was adopted for cluster.^19^ Hamming distance (Equation 1) was chosen as the dissimilarity metric, as it is naturally suited to binary feature spaces: it does not assume continuous data and assigns equal weight to the presence and absence of each clinical feature. In Equation 1, *xi* and *xj* are two samples, *p* is the total number of features, and *l* denotes the feature index. The indicator function **1**[*xil* ≠ *xjl*] equals 1 when the values differ between the two samples. The silhouette coefficient (Equation 2) was used to select the optimal number of clusters, where *a*(*i*) represents the average intra-cluster distance and *b*(*i*) the minimum average inter-cluster distance. The term max{*a*(*i*), *b*(*i*)}is used for normalization, constraining the silhouette coefficient to the interval [−1, 1].

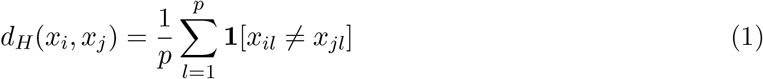

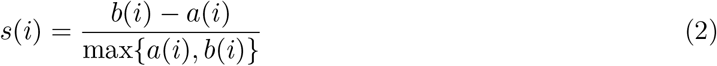

As a density-based clustering algorithm, DBSCAN groups observations in high-density regions without requiring a predefined number of clusters,^20^ offering robustness to outliers and the ability to detect irregularly shaped groups. The algorithm requires two parameters: *ε*, defining the neighbour-hood radius, and MinPts, the minimum number of samples required to form a dense region. The k-NN distance plot,^20^ computed for *k* ∈ {3, 5, 7}, was used to guide the selection of *ε*, informing a search range of *ε* ∈ {0.09, 0.10, 0.11, 0.12}. *ε* and MinPts∈ {3, 5, 7, 10, 15}were then jointly selected via a systematic grid search, choosing the configuration that maximised the number of clusters while keeping the noise proportion low, as the goal was to separate individuals into distinct phenotypic subgroups.

Finally, Latent Class Analysis (LCA) is a probabilistic clustering method widely used in medical research for identifying subphenotypes and uncovering latent group structures within clinical datasets.^21, 26^ Rather than assigning each individual to a single cluster, LCA estimates the posterior probability of membership across all latent classes.

LCA requires the number of classes k as an input parameter. The optimal value of k was determined using model selection criteria, specifically the Akaike Information Criterion (AIC)(Equation 3) and the Bayesian Information Criterion (BIC)(Equation 4). In these formulations, *L* denotes the maximized log-likelihood of the model, *p* represents the number of free parameters, *n* is the sample size, and *K* corresponds to the number of latent classes. Lower AIC and BIC values indicate better model fit, to balance model interpretability and cluster granularity while avoiding overfitting, both criteria were jointly considered in the selection of the optimal number of classes since BIC tends to avoid complexity and AIC to maximise it. Relative entropy was additionally examined as a measure of classification certainty, with values closer to 1 indicating clearer separation of individuals into latent classes (Equation 5), where *N* is the total number of individuals, *K* the number of latent classes, and *pik* the posterior probability of individual *i* belonging to class *k*.

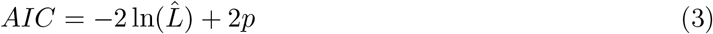

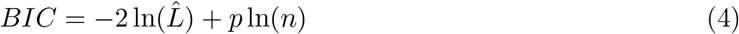

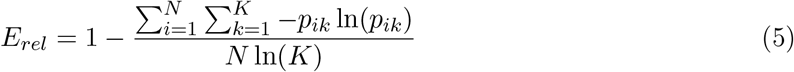

### 2.5 Clustering Framework Comparison and Visualisation

Based on these metrics, the best parameter configuration was selected by considering both statistical performance and the interpretability of the resulting clusters. To compare clustering approaches, we used the silhouette score (Equation 2) with Hamming distance (Equation 1) and Multiple Correspondence Analysis (MCA) for cluster visualization. The goal was to identify the configuration with the highest silhouette score and the most visually separated and interpretable clusters.

Multiple Correspondence Analysis (MCA) is a dimensionality reduction technique designed for categorical and binary data,^27^ projecting observations into a lower-dimensional space while preserving the main variance structure. In this study, MCA was applied to visualise cluster separability by projecting individuals into a two-dimensional space, where similar clinical profiles appear closer together and overlapping groups become visually apparent.

### 2.6 Association Rules

Based on the best clustering technique, each cluster was treated as an independent dataset for association rule extraction. Association rules identify combinations of clinical features that frequently co-occur, expressing the likelihood that the presence of a set of symptoms is associated with a given outcome. The Apriori algorithm^28^ was applied to all individuals within each cluster, including both deletion-positive and deletion-negative cases, with the consequent fixed as confirmed deletion status and the number of antecedents limited to four to ensure interpretability. A minimum confidence of 0.8 and minimum support of 0.04 were defined, where confidence measures the conditional probability of the consequent given the antecedent, and support reflects the frequency with which a rule appears in the dataset. Given the heterogeneity of individuals, support values were empirically determined to balance the number of generated rules while avoiding extremely rare patterns. The top 10 rules per cluster were selected based on lift (Equation 6), defined as the ratio of rule confidence to the marginal support of the consequent — where A denotes the set of antecedent symptoms, C the consequent (deletion-positive status), and lift *>* 1 indicates that the co-occurrence is more frequent than expected by chance. For instance, a rule may state that the co-occurrence of Long face, absence of Tetralogy of Fallot, Intellectual disability, and Velopharyngeal insufficiency is associated with a confirmed deletion.

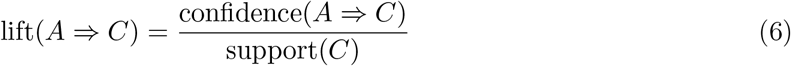

## 3 Results

The proposed pipeline identified four distinct groups within the 22q11.2 deletion syndrome cohort via LCA, which achieved the highest cluster separability among the three evaluated frameworks (silhouette = 0.165). MCA projections (Fig. 3) corroborate this finding, showing clearer group separation under LCA, although similar results for k-medoids.

K-medoids resulted in relatively low silhouette scores, ranging from 0.070 to 0.126. Although k=4 achieved the highest silhouette value, indicating the best separation among the evaluated configurations, the overall scores remained low. This suggests limited cluster compactness, indicating a possible overlap between clusters.

DBSCAN yielded the weakest performance among the three evaluated frameworks. Comparison across parameter combinations showed that DBSCAN frequently collapsed observations into a single dominant cluster under lower-noise configurations, limiting its utility for detailed phenotypic profiling. The best identified configuration, *ε*=0.09 and min_samples=3, produced three clusters but resulted in 79 noise samples (30% of the cohort), with two markedly small clusters and the majority of observations concentrated in a single group, as illustrated in Fig. 3.

LCA results indicated that k=4 provided the optimal balance between statistical fit and clinical interpretability, assessed through the degree of cluster separation and the coherence of the resulting subgroup profiles with established medical knowledge. While the 3-cluster solution achieved the lowest BIC (AIC = 10492.30, BIC = 11151.74), the difference relative to the 4-cluster solution (AIC = 10334.11, BIC = 11214.54) was small (ΔBIC = 62.8). Although BIC applies a stronger penalty for model complexity and marginally favoured the simpler solution, the 4-cluster model was preferred as it produced more granular and distinct combinations of features. AIC continued to decrease with increasing k, suggesting overfitting beyond k=4 when considered alongside BIC. The 4-class solution yielded a relative entropy of 0.987, comparable to the highest value observed (*Erel*=0.989 at K=6), indicating high classification certainty despite lower model complexity.

### 3.1 Cluster Characterisation

Symptom prevalence was computed as the proportion of individuals presenting each clinical sign within each cluster, as illustrated in Fig. 4.

Cluster 0 (C0) comprised 11 cases with a confirmed deletion, representing 14% of the cluster. It was almost exclusively defined by congenital heart defects (98%), with approximately 80% of individuals also presenting facial signs, including Tetralogy of Fallot (46%), bulbous nose (44%), and hooded eyelids (41%). Notably, orofacial cleft was completely absent in this cluster despite being present in all others, as was velopharyngeal insufficiency, and schizophrenia, features that emerge in other subgroups.

Cluster 1 (C1) represented the largest positive subgroup, comprising 32 cases with a confirmed deletion (49% of the cluster). While sharing the cardiac involvement observed in Cluster 0 as congenital heart defects were present in 86% of cases, this cluster was distinctly characterised by the co-occurrence of structural orofacial anomalies — most notably orofacial cleft (72%), which was completely absent in Cluster 0. Additional predominant features included facial signs (60%) and alar hypoplasia (40%), suggesting a craniofacial-palatal phenotype combined the cardiac profile.

Cluster 2 (C2) diverged markedly from the preceding groups, representing the most neurode-velopmentally distinct profile in the cohort with 19 cases with a confirmed deletion. Its defining feature was intellectual disability, present in 89% of cases with a confirmed deletion, a prevalence observed in no other cluster at comparable levels. Congenital heart defects were substantially lower (32%) and Tetralogy of Fallot nearly absent (3%), in contrast to Clusters 0 and 1. This cluster also recorded the highest prevalence across all groups for orofacial cleft, hypernasal speech, and learning disability, consolidating a craniofacial-neurodevelopmental phenotype largely separated from cardiac involvement.

Cluster 3 (C3) comprised 20 individuals with the deletion, representing 32% of the subgroup, and was distinguished by a markedly lower prevalence of congenital heart defects (3%) and Intellectual disability (6%), both common in Cluster 2, alongside a more heterogeneous overall profile, with no single feature exceeding 80%. Its predominant characteristics were other facial signs (63%), learning disabilities (44%), orofacial cleft (47%), and other psychiatric abnormalities (44%).

Following cluster identification, individuals with the deletion were isolated within each subgroup to examine symptom prevalence patterns exclusively among confirmed cases, independent of the individuals without the deletion present in each cluster. Congenital heart defect was absent in Cluster 3, while intellectual disability emerged as a strong and near-exclusive marker of Cluster 2, with negligible presence in Clusters 0 and 1. Although facial signs were present across all clusters, orofacial cleft was entirely absent in Cluster 0. Tetralogy of Fallot appeared only in Clusters 0 and 1, whereas ventricular septal defect and pulmonary atresia were prominent in Cluster 0, and interrupted aortic arch was restricted to Clusters 0 and 1. Thymic aplasia was notably concentrated in Cluster 0.

Cluster 1 showed a distinctive profile, being the only group to present small mouth, truncus arteriosus, hypoparathyroidism, renal abnormality, and bifid uvula. Urethral abnormality was exclusive to Cluster 3, and T-cell abnormality appeared solely in Cluster 2. Psychiatric abnormalities, including schizophrenia, were more prevalent in Clusters 2 and 3. Notably, a family history of palatal anomaly was found exclusively in Cluster 2, which also presented the highest concentration of facial anomalies. Learning disability was shared across all clusters, suggesting it represents a common baseline manifestation rather than a cluster-defining characteristic.

### 3.2 Association Rule Mining per Cluster

Following the identification of distinct subgroups via unsupervised clustering, association rule mining was applied to characterise the clinical profiles of individuals with a confirmed deletion within each cluster, restricting the consequent to confirmed deletion status. Each rule takes the form of a set of co-occurring clinical features (antecedents) associated with a confirmed deletion (consequent). For instance: *Hooded eyelids (Yes), Velopharyngeal insufficiency (Yes), Global developmental delay (Yes)* ⇒ *Deletion positive (Yes)*, indicating that the co-occurrence of these three features is associated with a confirmed 22q11.2 deletion within that subgroup. Table 1 summarises the clinical features present across the top 10 association rules per cluster, selected by lift; the full rules including antecedent combinations and metrics are provided in the Supplementary Material.

**Table 1:** Clinical Features Present in the Highest-Lift Association Rules per Cluster.

| **Clinical feature** | **C0** | **C1** | **C2** | **C3** |
| *Mean lift* | *5.89* | *2.03* | *1.95* | *3.30* |
|---|---|---|---|---|
| Other facial signs | ✓ | ✓ | ✓ | ✓ |
| Ventricular septal defect (VSD) | ✓ | ✓ |  |  |
| Congenital heart defect | ✓ | ✓ |  |  |
| Learning disability |  | ✓ | ✓ | ✓ |
| Orofacial cleft |  | ✓ | ✓ | ✓ |
| Long face |  |  | ✓ | ✓ |
| Velopharyngeal insufficiency |  |  | ✓ | ✓ |
| Hooded eyelids |  | ✓ | ✓ |  |
| Bulbous nose |  | ✓ |  |  |
| Prominent nasal bridge |  | ✓ |  |  |
| High nasal bridge |  | ✓ |  |  |
| Atrial septal defect (ASD) |  | ✓ |  |  |
| Hypernasal speech |  |  | ✓ |  |
| Intellectual disability |  |  | ✓ |  |
| Global developmental delay |  |  | ✓ |  |
| Thymic abnormality |  |  | ✓ |  |
| Tubular nose |  |  | ✓ |  |
| Alar hypoplasia |  |  |  | ✓ |

In Cluster 0, the identified rules reached a lift of 5.8, reflecting a markedly non-random co-occurrence of these clinical features within this cluster. The predominant associations involved ventricular septal defect, congenital heart defect, and other facial signs. This subgroup yielded only four association rules meeting the defined thresholds, despite the selection criteria allowing up to ten rules per cluster.

In Cluster 1, association rules reached lifts approaching 2.0, indicating that the associated clinical features co-occurred at approximately twice the frequency expected under statistical independence within the cluster. The rules show that the presence of isolated atrial septal defect already represented a strong diagnostic suspicion. Furthermore, combinations involving ventricular septal defect associated with craniofacial and neurodevelopmental manifestations also demonstrated high discriminative power. Overall, the rules suggest the existence of two main phenotypic profiles within this cluster. The first corresponds to a classic cardiofacial phenotype, characterised by ventricular septal defect, orofacial cleft, learning disability, and congenital heart defect. The second represents a predominantly facial phenotype, marked by bulbous nose, prominent nasal bridge, high nasal bridge, and the presence of hooded eyelids.

In Cluster 2, association rules achieved lifts approaching 2.0, indicating that the associated clinical features co-occurred at nearly twice the frequency expected under statistical independence within the cluster. This subgroup was characterised by a predominantly craniofacial and velopharyngeal phenotype, marked by long face, hypernasal speech, velopharyngeal insufficiency, orofacial cleft, and global developmental delay. Notably, the combination of long face, velopharyngeal insufficiency, and absence of Tetralogy of Fallot was sufficient to exclusively identify individuals with a confirmed deletion, highlighting palatal and speech features as strong phenotypic markers. Additional patterns involving hypernasal speech, orofacial cleft, and global developmental delay further reinforced the relevance of velopharyngeal and craniofacial manifestations in this subgroup. Cluster-specific facial features, including tubular nose and hooded eyelids, were also associated with positive diagnosis in the absence of severe congenital heart defects, consolidating a predominantly craniofacial and neurodevelopmental profile.

In Cluster 3, association rules yielded lifts up to 3.9, indicating that the associated clinical features co-occurred at nearly four times the frequency expected under statistical independence. This subgroup represents a predominantly craniofacial and neurodevelopmental phenotype, characterised by learning disability, velopharyngeal insufficiency, long face, and orofacial cleft. Notably, the combination of velopharyngeal insufficiency and learning disability demonstrated strong discriminative power for positive diagnosis even in the absence of congenital heart defects. Additional rules suggested an association between orofacial cleft and global developmental delay.

## 4 Discussion

The diagnostic journey of 22q11.2 deletion syndrome is frequently marked by prolonged referral across multiple specialists before a correct diagnosis is established. To optimise cost and referral decisions, a hierarchical approach has been adopted within Brazil’s Craniofacial Project (BCFP) partner institutions: clinical screening filters individuals first, followed by low-cost genetic tests, with CMA reserved for cases remaining under suspicion.^18^ However, the screening protocol evaluates clinical signs independently, without accounting for symptom co-occurrence — which, as this study demonstrates, differs meaningfully across subgroups, leaving individuals with mild or atypical presentations at risk of being missed.

This study employed Latent Class Analysis to identify subgroups based on the clinical signs of 22q11.2 deletion syndrome, further characterised through symptom prevalence analysis and association rule mining to uncover co-occurring patterns among individuals with a confirmed deletion. Four phenotypically distinct clusters were identified: Cardiac-predominant (C0), Craniofacial-palatal (C1), Neurodevelopmental-psychiatric (C2), and Mild-craniofacial (C3). A certain degree of overlap between subgroups was anticipated given the well-documented clinical heterogeneity of the condition,^8^ and was evidenced by the low overall silhouette scores observed for both LCA and K-medoids, as well as the limited cluster separation visible in the MCA projections. Previous studies applying clustering to cognitive profiles in 22q11.2 have reported continuous gradations rather than discrete groups;^17^ however, by examining a broader set of clinical features spanning cardiac, craniofacial, immunological, and neurodevelopmental domains, this study captures a wider spectrum of phenotypic variation. While some overlap between subgroups is expected, the concentration of cardiac features in C0 and neurodevelopmental features in C2 potentially suggests that incorporating a broader spectrum of clinical characteristics across multiple domains may yield more distinct subgroup separation when applying clustering methods to individuals with suspected deletion. The following clinical interpretation reflects the integration of data-driven findings with the assessment of medical specialists in clinical genetics and craniofacial anomalies.

Cluster 0, characterised by a predominance of congenital heart defects, highlights the importance of considering 22q11.2 deletion as a differential diagnosis in cardiac presentations; however, the relatively low deletion-positive rate within this subgroup corroborates existing evidence that isolated congenital heart disease alone is an insufficient predictor of deletion status.^29^ Notably, the higher positivity observed in cases of ventricular septal defect within this cluster complements prior descriptions in the literature,^18, 30^ while also revealing that this association is not yet adequately reflected in the current screening protocol^18^ — suggesting a concrete opportunity for targeted refinement.

In the profile drawn by Cluster 1, cardiac malformations showed their relevance in the investigation of suspected cases of 22q11.2 deletion when associated with characteristic craniofacial anomalies and dysmorphisms (bulbous nose, prominent nasal bridge, high nasal bridge and presence of hooded eyelids). This result is of great relevance in pointing to a high positivity rate when associating cardiac defects with common craniofacial alterations, such as orofacial cleft and dysmorphisms, which may not be analyzed together during the diagnostic odyssey. Thus, this cluster outlines an important clinical parameter in characterising suspected cases of 22q11.2 deletion, defining a relevant phenotypic profile to be disseminated across various health care settings. In addition, although the cluster reinforces the importance of the presence of several signs in cases of positive suspicion for the syndrome, the pattern found reveals a positive trend in cases of isolated atrial septal defect and in the absence of conotruncal defects, a type of alteration already associated with a higher prevalence of 22q11.2 deletion.^31^

Cluster 2 highlighted the relevance of suspecting 22q11.2 deletion in cases of craniofacial anomalies, such as orofacial cleft and velopharyngeal insufficiency, especially when associated with global developmental delay. The importance of diagnostic investigation in cases of orofacial cleft was already evident in the literature, since 22q11.2 deletion is the most common genetic cause for these alterations.^7^ However, the present analysis shows an important association of positive cases with the concomitant presence of global developmental delay and learning disability. Notably, these findings reinforce that the absence of congenital heart disease should not reduce clinical suspicion, as the craniofacial and neurodevelopmental profiles identified across clusters represent strong, literature-supported diagnostic indicators of the 22q11.2 deletion in their own right.^32^ In addition, in cases of hypernasal speech, velopharyngeal insufficiency, long face associated with developmental deficit, the absence of Tetralogy of Fallot proved to be a good differential for positive cases, corroborating the distinct phenotypic profile already presented in the literature and compatible with another cluster.^31^ Cluster 3 was the most heterogeneous among the groups, with a low prevalence of specific characteristics and focusing the analysis on facial signs and anomaly.

When comparing individuals who meet the screening criteria proposed by Monteiro et al.,^18^ across cluster profiles, the protocol showed varying levels of sensitivity and specificity, as indicated in Table 2. Sensitivity reflects the protocol’s ability to correctly identify true individuals with a confirmed deletion, minimising missed diagnoses, while specificity reflects its capacity to avoid unnecessary referrals of individuals who do not carry the deletion. C0 demonstrated the most balanced performance (sensitivity 72.7%, specificity 72.9%), indicating that the protocol is best suited for cardiac-predominant presentations. C1 and C2 presented low specificity (25.0% and 27.8%) but relatively higher sensitivity (75.0% and 57.9%), meaning that while most positive cases are captured, a substantial proportion of unaffected individuals are unnecessarily referred for genetic testing — contributing disproportionately to testing costs. In contrast, C3 showed higher specificity (58.3%) but the lowest sensitivity (50.0%), suggesting that half of the individuals with a confirmed deletion who present with this phenotypic profile may be missed by current screening criteria.

**Table 2:** Screening Protocol Performance per Cluster.

| **Cluster** | **Sensitivity (%)** | **Specificity (%)** |
|---|---|---|
| C0 — Cardiac-predominant | 72.73 | 72.86 |
| C1 — Craniofacial-palatal | 75.00 | 25.00 |
| C2 — Neurodevelopmental-psych. | 57.89 | 27.78 |
| C3 — Mild-craniofacial | 50.00 | 58.33 |

Clusters C1 and C2, which presented the lowest specificity, are likely the primary drivers of unnecessary genetic referrals, directing a substantial proportion of unaffected individuals toward costly confirmatory testing. Conversely, individuals in C2 and C3, with sensitivities of 57.9% and 50.0%, face a higher risk of remaining undiagnosed — circulating across multiple specialists without receiving a confirmed 22q11.2 diagnosis, perpetuating the diagnostic odyssey described in the literature.^33^ The Neurodevelopmental-psychiatric subgroup (C2) presents the most critical challenge, combining low sensitivity and low specificity — meaning that individuals with a confirmed deletion who present with this profile are simultaneously at risk of being missed by current screening criteria and of triggering unnecessary referrals among individuals without the deletion who exhibit overlapping neurodevelopmental presentations. This may occur since these manifestations are also common across a broad range of neurodevelopmental and psychiatric conditions, limiting their specificity for syndrome recognition.^34^

In future work, these findings may support the integration of symptom co-occurrence patterns as an additional scoring criterion within the existing protocol, building upon established approaches to phenotype-driven clinical decision support for rare disease diagnosis.^18^ Individuals whose clinical profiles match the co-occurrence patterns identified in deletion-positive subgroups could receive greater diagnostic weight even when individual symptoms do not meet referral thresholds — effectively transforming the current rule-based scoring system into a pattern-aware referral mechanism. This may improve early identification of 22q11.2 deletion syndrome, particularly for phenotypic profiles currently underserved by uniform screening criteria, and represents a tractable direction for extending the pipeline toward a clinically deployable decision support tool.

Several data-related limitations should be acknowledged. The unbalanced dataset may bias the results, and focusing only on positive symptoms limited the identification of negative patterns. One-hot encoding increased data dimensionality, potentially overweighting some clinical variables during clustering. Additionally, keeping features with overlapping definitions may have introduced collinearity and redundancy, a trade-off accepted to preserve clinical relevance and avoid further sample loss due to incomplete data in other variables.

These results come from a single Brazilian reference database, and the specific subgroups and symptom patterns may differ in other populations and healthcare settings. The pipeline itself, however, is not tied to this cohort and can be applied to other databases, although the phenotypic profiles it identifies may vary. Replication in independent cohorts is needed.

## 5 Conclusion

The present study proposed a five-stage unsupervised pipeline for the phenotypic stratification of 22q11.2 deletion syndrome, a rare condition presenting marked clinical heterogeneity. LCA achieved the highest cluster separability (silhouette = 0.165), identifying four phenotypically distinct sub-groups: Cardiac-predominant (n=11), Craniofacial-palatal (n=32), Neurodevelopmental-psychiatric (n=19), and Mild-craniofacial (n=20). Assessment of the current screening protocol revealed ad-equate overall sensitivity; however, sensitivity was markedly reduced in the Neurodevelopmental-psychiatric and Mild-craniofacial subgroups (57.9% and 50.0%, respectively), indicating that a sub-stantial proportion of individuals with a confirmed deletion within these profiles may be missed by current referral criteria. Validated by medical specialists, these findings suggest that phenotypic stratification may support more targeted referral pathways, reducing unnecessary genetic examinaions and improving the cost-effectiveness of diagnostic screening for 22q11.2 deletion syndrome.

## Data Availability

Data may be available upon reasonable request to the coordinating center, subject to institutional and ethical approval.

