## Supplementary Material for "Unsupervised Phenotypic Stratification of 22q11.2 Deletion Syndrome for Improved Clinical Screening"

This supplementary material provides additional details supporting the methods and results reported in the main manuscript. It includes: the distribution of positive, negative, and missing responses across all clinical features in the total cohort; the parameter selection procedures for DBSCAN and LCA; a detailed symptom prevalence characterisation of each cluster restricted to confirmed deletion-positive individuals; and the complete set of association rules extracted per cluster.

The supplementary material is organised as follows: Section S1 describes the dataset and feature details; Section S2 presents the parameter selection procedures for K-medoids and DBSCAN; Section S3 reports the model selection process for Latent Class Analysis (LCA); Section S4 provides the cluster characterisation restricted to confirmed positive individuals; and Section S5 presents the complete set of association rules extracted per cluster.

#### S1. Dataset and Feature Details

This section presents the symptom prevalence distribution across the full cohort prior to outlier removal, covering the 47 clinical attributes selected for analysis. Each feature is shown in Fig.1 with the proportion of patients recorded as present, absent, or missing.

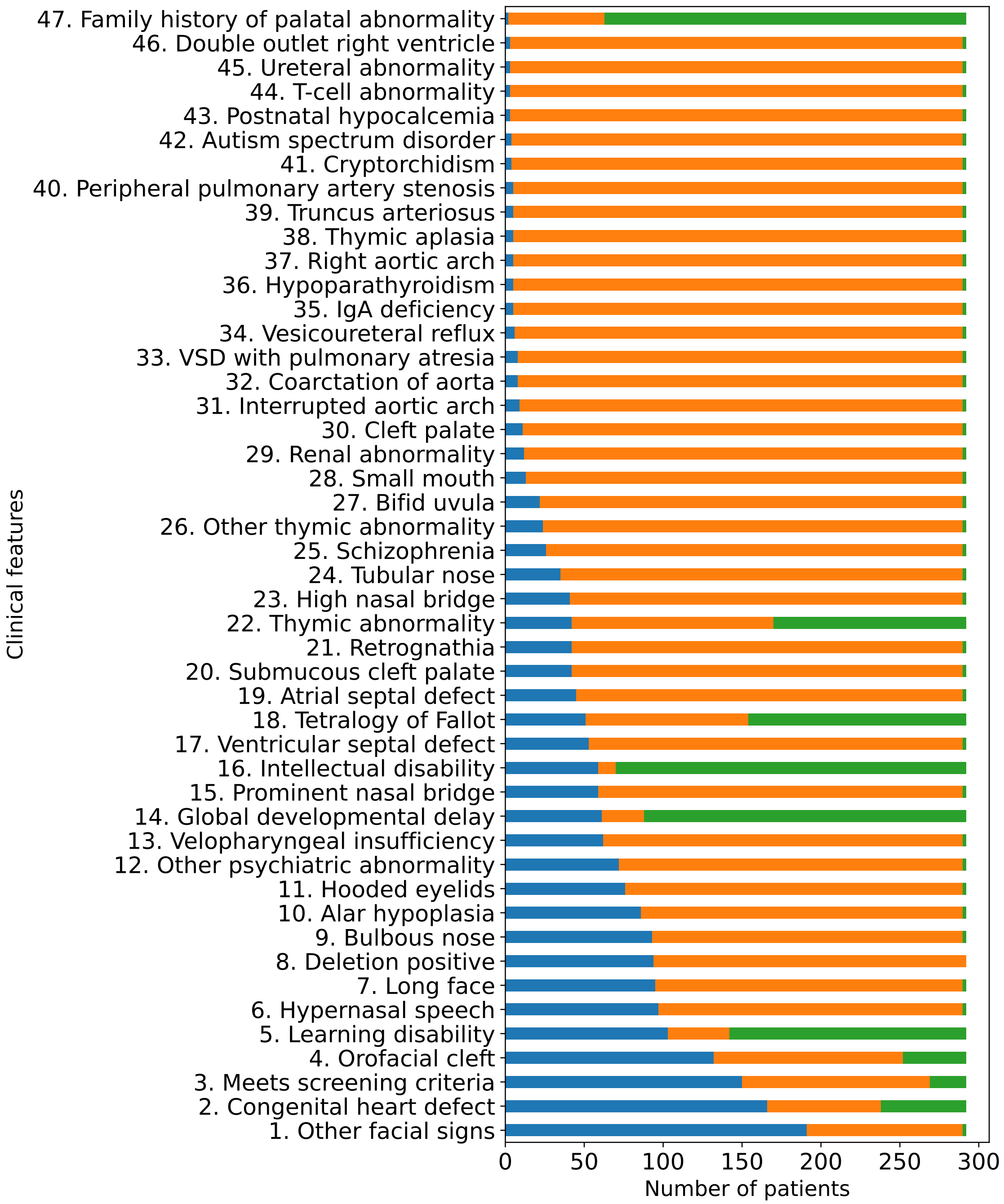


Figure 1: Distribution of the 47 clinical features across the cohort. Blue bars represent positive cases, orange bars represent negative cases, and green bars indicate missing values.

### S2. K-medoids Parameter Selection

K-medoids clustering was evaluated for $k \in \{2, \ldots, 7\}$ using Hamming distance with the `alternate` optimisation method [1]. Silhouette scores remained consistently low across all configurations (range: 0.070–0.126), with $k = 4$ achieving the highest value (0.126), as indicated in Table 1.

Table 1: K-medoids Silhouette Scores by Number of Clusters

| $k$ | **Silhouette Score** |
|---|---|
| 2 | 0.100 |
| 3 | 0.115 |
| 4 | **0.126** |
| 5 | 0.119 |
| 6 | 0.111 |
| 7 | 0.109 |
| 8 | 0.071 |
| 9 | 0.070 |

**Bold**: Best configuration

### S3. DBSCAN Parameter Selection

The k-nearest neighbour distance plot was computed for $k \in \{3, 5, 7\}$ to assess the density structure of the data prior to parameter selection [2]. Across all evaluated values of $k$, the resulting curves show an approximately linear increase in Hamming distance, with no clearly identifiable elbow point, suggesting the absence of a density structure in the data. This result was used to inform the range of values evaluated in the subsequent grid search.

A systematic grid search, shown in Tab.2, was conducted over combinations of $\varepsilon \in \{0.09, 0.10, 0.11, 0.12\}$ and $MinPts \in \{3, 5, 7, 10, 15\}$, evaluating the resulting number of clusters and proportion of noise samples for each configuration (Table 2). The configuration maximising cluster count while minimising noise ($\varepsilon = 0.09$, $MinPts = 3$).

Table 2: DBSCAN Parameter Grid Search Results

| $\varepsilon$ | $MinPts$ | **Clusters** | **Noise** | **Noise (%)** |
|---|---|---|---|---|
| **0.09** | **3** | **3** | **79** | **30** |
| 0.10 | 3 | 2 | 51 | 20 |
| 0.11 | 3 | 2 | 51 | 20 |
| 0.12 | 3 | 1 | 20 | 8 |
| 0.12 | 5 | 1 | 24 | 9 |
| 0.12 | 7 | 1 | 27 | 10 |
| 0.12 | 10 | 1 | 38 | 15 |
| 0.12 | 15 | 1 | 53 | 20 |
| 0.10 | 5 | 1 | 65 | 25 |
| 0.11 | 5 | 1 | 65 | 25 |
| 0.10 | 7 | 1 | 72 | 28 |
| 0.11 | 7 | 1 | 72 | 28 |
| 0.10 | 10 | 1 | 84 | 32 |
| 0.11 | 10 | 1 | 84 | 32 |
| 0.10 | 15 | 1 | 99 | 38 |
| 0.11 | 15 | 1 | 99 | 38 |
| 0.09 | 5 | 1 | 105 | 40 |
| 0.09 | 7 | 1 | 122 | 47 |
| 0.09 | 10 | 1 | 146 | 56 |
| 0.09 | 15 | 1 | 168 | 64 |

**Bold**: Best configuration

### S4. LCA Model Selection

Latent Class Analysis models were fitted for $K \in \{2, 3, 4, 5, 6, 7\}$ latent classes, shown in Table 3. Model selection was based on AIC and BIC [3]. While AIC decreased monotonically, BIC reached its minimum at $K = 3$ (BIC = 11,151.74).

Table 3: LCA model selection criteria and classification certainty for candidate numbers of latent classes ($K$).

| $K$ | AIC | BIC | $E_{\text{rel}}$ |
|---|---|---|---|
| 2 | 10785.67 | 11224.11 | 0.954 |
| 3 | 10492.31 | **11151.74** | 0.962 |
| 4* | 10334.11 | 11214.54 | 0.987 |
| 5 | 10185.68 | 11287.12 | 0.972 |
| 6 | 10118.66 | 11441.09 | **0.989** |
| 7 | **10085.52** | 11628.95 | 0.984 |

**Bold**: best value in each column. *Selected model.

### S5. LCA Model Selection

### S5. Full Association Rules per Cluster

Association rules were extracted for each cluster independently, restricted to confirmed positive individuals. The following tables present all rules meeting the defined thresholds (minimum confidence = 0.8, minimum support = 0.04), ranked by lift. The main manuscript reports the top rules per cluster; complete results are provided in Tables4, 5, 6, 7.

Table 4: Association Rules – Cluster 0

| Antecedents | Consequents | Support | Confidence | Lift |
|---|---|---|---|---|
| Congenital heart defect (Yes), Ventricular septal defect (Yes), Family history of palatal abnormality (No), Other facial signs (Yes) | Deletion positive (Yes) | 0.05 | 0.80 | 5.89 |
| Ventricular septal defect (Yes), Family history of palatal abnormality (No), Other facial signs (Yes) | Deletion positive (Yes) | 0.05 | 0.80 | 5.89 |
| Other facial signs (Yes), Ventricular septal defect (Yes), Intellectual disability (Missing), Family history of palatal abnormality (No) | Deletion positive (Yes) | 0.05 | 0.80 | 5.89 |
| Other facial signs (Yes), Ventricular septal defect (Yes), Orofacial cleft (No), Family history of palatal abnormality (No) | Deletion positive (Yes) | 0.05 | 0.80 | 5.89 |

Table 5: Association Rules – Cluster 1

| **Antecedents** | **Consequents** | **Support** | **Confidence** | **Lift** |
|---|---|---|---|---|
| Learning disability (Missing), Global developmental delay (Missing), Atrial septal defect (Yes), Tetralogy of Fallot (Missing) | Deletion positive (Yes) | 0.06 | 1.00 | 2.03 |
| Ventricular septal defect (Yes), Tetralogy of Fallot (No), Learning disability (Yes), Orofacial cleft (Yes) | Deletion positive (Yes) | 0.05 | 1.00 | 2.03 |
| Hooded eyelids (Yes), Intellectual disability (Missing), Congenital heart defect (Yes), Tetralogy of Fallot (Missing) | Deletion positive (Yes) | 0.05 | 1.00 | 2.03 |
| Intellectual disability (Missing), Atrial septal defect (Yes), Bulbous nose (Yes), Prominent nasal bridge (Yes) | Deletion positive (Yes) | 0.05 | 1.00 | 2.03 |
| Thymic abnormality (Missing), Bulbous nose (Yes), Prominent nasal bridge (Yes) | Deletion positive (Yes) | 0.06 | 1.00 | 2.03 |
| Congenital heart defect (Yes), Family history of palatal abnormality (Missing), Other facial signs (Yes), High nasal bridge (Yes) | Deletion positive (Yes) | 0.05 | 1.00 | 2.03 |
| High nasal bridge (Yes), Other facial signs (Yes), Bulbous nose (Yes) | Deletion positive (Yes) | 0.05 | 1.00 | 2.03 |
| Global developmental delay (Missing), Atrial septal defect (Yes), Bulbous nose (Yes), Prominent nasal bridge (Yes) | Deletion positive (Yes) | 0.05 | 1.00 | 2.03 |
| Thymic abnormality (Missing), Tetralogy of Fallot (No), Learning disability (Missing), Bulbous nose (Yes) | Deletion positive (Yes) | 0.08 | 1.00 | 2.03 |
| Intellectual disability (Missing), Thymic abnormality (Missing), Learning disability (Missing), Prominent nasal bridge (Yes) | Deletion positive (Yes) | 0.06 | 1.00 | 2.03 |

Table 6: Association Rules – Cluster 2

| **Antecedents** | **Consequents** | **Support** | **Confidence** | **Lift** |
|---|---|---|---|---|
| Long face (Yes), Tetralogy of Fallot (No), Intellectual disability (Yes), Velopharyngeal insufficiency (Yes) | Deletion positive (Yes) | 0.11 | 1.00 | 1.95 |
| Long face (Yes), Learning disability (Yes), Family history of palatal abnormality (No), Global developmental delay (Yes) | Deletion positive (Yes) | 0.05 | 1.00 | 1.95 |
| Family history of palatal abnormality (Missing), Hypernasal speech (Yes), Thymic abnormality (Yes), Global developmental delay (Missing) | Deletion positive (Yes) | 0.05 | 1.00 | 1.95 |
| Global developmental delay (Missing), Thymic abnormality (Yes), Orofacial cleft (Yes) | Deletion positive (Yes) | 0.05 | 1.00 | 1.95 |
| Family history of palatal abnormality (Missing), Hypernasal speech (Yes), Intellectual disability (Yes), Thymic abnormality (Yes) | Deletion positive (Yes) | 0.05 | 1.00 | 1.95 |
| Thymic abnormality (No), Global developmental delay (Missing), Hypernasal speech (Yes), Family history of palatal abnormality (No) | Deletion positive (Yes) | 0.05 | 1.00 | 1.95 |
| Long face (Yes), Tetralogy of Fallot (No), Velopharyngeal insufficiency (Yes), Other facial signs (Yes) | Deletion positive (Yes) | 0.05 | 1.00 | 1.95 |
| Long face (Yes), Thymic abnormality (Missing), Learning disability (Yes), Family history of palatal abnormality (No) | Deletion positive (Yes) | 0.05 | 1.00 | 1.95 |
| Congenital heart defect (No), Tubular nose (Yes), Tetralogy of Fallot (Missing), Hypernasal speech (Yes) | Deletion positive (Yes) | 0.05 | 1.00 | 1.95 |
| Hooded eyelids (Yes), Velopharyngeal insufficiency (Yes), Global developmental delay (Yes) | Deletion positive (Yes) | 0.05 | 1.00 | 1.95 |

Table 7: Association Rules – Cluster 3

| **Antecedents** | **Consequents** | **Support** | **Confidence** | **Lift** |
|---|---|---|---|---|
| Congenital heart defect (No), Learning disability (Yes), Velopharyngeal insufficiency (Yes), Global developmental delay (Missing) | Deletion positive (Yes) | 0.05 | 1.00 | 3.90 |
| Long face (Yes), Global developmental delay (Missing), Learning disability (Yes), Velopharyngeal insufficiency (Yes) | Deletion positive (Yes) | 0.05 | 1.00 | 3.90 |
| Global developmental delay (Missing), Learning disability (Yes), Long face (Yes), Orofacial cleft (Yes) | Deletion positive (Yes) | 0.06 | 0.83 | 3.25 |
| Congenital heart defect (No), Learning disability (Yes), Other facial signs (Yes), Thymic abnormality (No) | Deletion positive (Yes) | 0.06 | 0.83 | 3.25 |
| Congenital heart defect (No), Learning disability (Yes), Velopharyngeal insufficiency (Yes) | Deletion positive (Yes) | 0.05 | 0.80 | 3.12 |
| Congenital heart defect (No), Learning disability (Yes), Velopharyngeal insufficiency (Yes), Orofacial cleft (Yes) | Deletion positive (Yes) | 0.05 | 0.80 | 3.12 |
| Congenital heart defect (No), Learning disability (Yes), Velopharyngeal insufficiency (Yes), Family history of palatal abnormality (Missing) | Deletion positive (Yes) | 0.05 | 0.80 | 3.12 |
| Congenital heart defect (No), Intellectual disability (Missing), Learning disability (Yes), Velopharyngeal insufficiency (Yes) | Deletion positive (Yes) | 0.05 | 0.80 | 3.12 |
| Long face (Yes), Learning disability (Yes), Velopharyngeal insufficiency (Yes) | Deletion positive (Yes) | 0.05 | 0.80 | 3.12 |
| Alar hypoplasia (Yes), Learning disability (Yes), Other facial signs (Yes), Thymic abnormality (No) | Deletion positive (Yes) | 0.05 | 0.80 | 3.12 |
